# A polygenic predictor of treatment-resistant depression using whole exome sequencing and genome-wide genotyping

**DOI:** 10.1101/19007161

**Authors:** Chiara Fabbri, Siegfried Kasper, Alexander Kautzky, Joseph Zohar, Daniel Souery, Stuart Montgomery, Diego Albani, Gianluigi Forloni, Panagiotis Ferentinos, Dan Rujescu, Julien Mendlewicz, Rudolf Uher, Cathryn M. Lewis, Alessandro Serretti

## Abstract

Treatment-resistant depression (TRD) occurs in ∼30% of patients with major depressive disorder (MDD) but the genetics of TRD was previously poorly investigated.

Whole exome sequencing and genome-wide genotyping were performed in 1320 MDD patients. Response to the first pharmacological treatment was compared to non-response to one treatment and non-response to two or more treatments (TRD). Differences in the risk of carrying damaging variants were tested. A score expressing the burden of variants in genes and pathways was calculated weighting each variant for its functional (Eigen) score and frequency, considering rare variants only and rare + common variants. Gene- and pathway-based scores were used to develop predictive models of TRD and non-response using gradient boosting in 70% of the sample (training) which were tested in the remaining 30% (testing), evaluating also the addition of clinical predictors. Independent replication was tested in STAR*D and GENDEP using exome array-based data.

After quality control 1209 subjects were included. TRD and non-responders did not show higher risk to carry damaging variants compared to responders. Genes/pathways associated with TRD included those modulating cell survival and proliferation, neurodegeneration and immune response. Significant prediction of TRD vs. response was observed in the testing sample which was improved by the addition of clinical factors. Some models were replicated, with a weaker prediction, in STAR*D and GENDEP when considering also clinical factors and in the extremes of the genetic score distribution.

These results suggested relevant biological mechanisms implicated in TRD and a new methodological approach to the prediction of TRD.

## 1. Introduction

Major depressive disorder (MDD) is the second leading cause of disability in middle-aged adults on a global scale (1). Despite the availability of a number of different pharmacological treatments, treatment-resistant depression (TRD) is estimated to occur in ∼30% of patients (2). TRD is usually defined as lack of response to at least two adequate treatments and it is associated with social and occupational impairment, suicidal thoughts, decline of physical health and increased health care utilization (3) (4). Annual costs for health care and lost productivity were estimated to be $5,481 and $4,048 higher, respectively, for a patient with TRD versus a patient with treatment-responsive depression (5).

In the future, biomarkers associated with TRD risk may contribute to improve the clinical management of MDD by providing an estimate of TRD genetic risk at baseline, by guiding the prescription of personalized treatments and the development of new drugs. Genetic variants are ideal biomarkers to predict treatment response and TRD: a genetic basis to treatment response has been demonstrated and genotyping can be performed in easily accessible samples with reasonable cost and time (6). The development of models able to predict the genetic risk of TRD at baseline would provide valuable information to personalize treatment prescription and hypothetically reduce the rate of TRD. Possible ways by which this could be achieved include: 1) identifying genetic predictors of non-response to specific antidepressant classes; 2) prescribing treatments with increased efficacy but limited availability because of costs constraints to patients having genetic risk for TRD. However, most existing pharmacogenomic studies were focused on measures of response to the last treatment without taking into account previous treatments, leaving the genetics of TRD largely unexplored (7). Another issue was the investigation of common variants only, while the possible role of rare variants was overlooked, despite they were suggested as one of the factors contributing to missing heritability of common traits (8). To the best of our knowledge, only a small pilot study (n=10) performed whole exome sequencing to the study of treatment response in MDD (but not TRD) and found that the bone morphogenetic protein (*BMP5*) gene may be associated with the therapeutic outcome (9).

The present study aimed to contribute in filling the existing gap in the knowledge of TRD genetics using whole exome sequencing and genome-wide genotyping to analyze the role of rare and common variants in the prediction of this phenotype and contribute to the development of predictive models potentially useful to personalize antidepressant prescription.

## 2. Patients and Methods

### 2.1. Sample

The Group for the Study of Resistant Depression (GSRD) sample was recruited within a multicenter, cross-sectional study including adult in- and outpatients with major depressive disorder (MDD) (DSM IV-TR criteria), as confirmed using the Mini International Neuropsychiatric Interview (MINI). Depressive symptom severity was assessed using the Montgomery and Åsberg Depression Rating Scale (MADRS) at study inclusion and at the onset of the current MDD episode. Information on previous and current antidepressant and other pharmacological treatments during the current MDD episode was collected as well as clinical-demographic characteristics. Antidepressant treatment was naturalistic according to best-clinical practice principles (Supplementary Table 1). The study protocol was approved by the local ethnic committees and the participant signed the written informed consent. Further details can be found elsewhere (10).

### 2.2. Phenotype, training and testing samples

TRD (treatment-resistant depression) was defined according to the most common definition of lack of response to at least two adequate antidepressant treatments (11), while non-response is referred to one treatment only. Adequate treatment was defined as an antidepressant treatment of minimum duration of 4 weeks at the minimum therapeutic dose according to drug labeling. Response was defined as a MADRS score < 22 and a score decrease of at least 50% compared to the onset of the current MDD episode. After quality control, the sample was split in a training (70%) and testing set (30%) which were balanced in terms of phenotypic distribution (TRD, non-response and response) using the partition function of groupdata2 R package, and they did not differ for gender, age, baseline depression severity or centre of recruitment.

### 2.3. Whole exome sequencing and genome-wide genotyping

Whole exome sequencing was performed using the Illumina HiSeq platform with 100 bp read length. Genome-wide genotyping was performed using the Illumina Infinium PsychArray 24 BeadChip (Illumina, Inc., San Diego) and these data were imputed as described in Supplementary Methods. Rare variants were extracted from exome sequence data and were defined as those having minor allele frequency (MAF) < 1/√(2*n*), where *n* is the sample size (12), which corresponded to 0.02 in GSRD. Information about DNA extraction, quality control of exome sequence data and genome-wide data are reported as Supplementary Methods. We compared the concordance of genotypes of SNPs available in both exome sequence and array data, splitting them in genotyped and imputed and by minor allele frequency (MAF). These comparisons were also relevant to determine the putative reliability of rare imputed variants in the replication samples. Subjects with discrepancies between genome-wide and exome sequence data were excluded (non-major homozygote genotype concordance ≤90% for rare variants and ≤95% for common variants).

### 2.4. Statistical analysis

#### 2.4.1. Variant annotation and distribution of functional variants

We tested if predicted detrimental/damaging variants obtained through exome sequencing were differently distributed between TRD patients, non-responders and responders. Variant annotation was performed using Variant Effect Predictor (Vep) release 90, using the –pick flag that chooses one block of annotation per variant, based on an ordered set of criteria (13). Annotations from SIFT, PolyPhen and functional consequence scores from the Sequence Ontology (SO) project were used to estimate the relative pathogenicity of variants (14) (15) (17). The use of scores which combine different variant annotations was also pursued and it is described in the next paragraph. The risk of carrying SIFT deleterious variants (scores<0.05), PolyPhen damaging or probably damaging variants (scores>0.45) and variants with SO functional score ≥ 0.90 and ≥ 0.70 in specific genes was compared across TRD patients, non-responders and responders using regression models adjusted for three population principal components and center of recruitment. Bonferroni correction was applied to account for multiple testing (the number of included genes was between 14,353 and 18,600 depending from the considered annotation). Additional details are reported as Supplementary Methods.

#### 2.4.2. Exome risk scores

These analyses aimed to estimate a weighted measure reflecting the burden of rare genetic variants exome-wide and in a gene- and pathway-based way. Secondly, we combined these measures with analogous estimations for common variants.

For rare variants, a score was calculated for each individual as 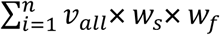 where *n* is the number of genetic variants within the considered unit (whole exome, gene or pathway), *v*_*all*_ is the number of alternative alleles, *w*_*s*_ is the corresponding functional score and *w*_*f*_ is the frequency weight for that variant. In this way, the score is not dependent from the presence of individual variants which could not be observed in some of the tested samples. A similar approach was previously applied to the study of schizophrenia risk using exome sequence data (18), but it was modified in this study by using different functional weighting (composite scores reflecting multiple annotations) and different frequency weighting (to allow the inclusion of rare but also common variants). Different sources for determining *w*_*s*_ were tested and compared (Eigen scores (19), CADD scores (16) and SO functional scores (15), see Supplementary Methods). The frequency weight was determined using a beta distribution based on the frequency of the alternative allele alt_all (*w*_*f*_ =dbeta(alt_all,1,25), according to the previous literature (12), see the corresponding curve in Supplementary Figure 1). Rare variants were extracted from exome sequence data as those with minor allele frequency (MAF < 1/√(2*n*), where *n* is the sample size (12), which corresponded to 0.02 in GSRD). Common intragenic variants were extracted from genome-wide genotyping data and clumped based on their functional scores *w*_*s*_ and linkage disequilibrium (LD) using Plink v.1.9 (Supplementary Methods). A smoother beta distribution was used to weight these variants based on frequency (*w*_*f*_ =dbeta(alt_all,0.5,0.5) (12), see curve in Supplementary Figure 1).

The obtained scores were tested for different distribution among the phenotypic groups considering rare variants only and the sum of the scores for rare and common variants. These tests were performed using regression models adjusted for three population principal components and centre of recruitment.

#### 2.4.3. Predictive modeling

Gene- and pathway-based scores (adjusted for the described confounders, more details in Supplementary Methods) were entered into a predictor selection process in the training sample using a five-fold cross-validation repeated 100 times for pathways and 20 times for genes, 500 and 100 rounds in total, respectively. In each round, one fifth of the training dataset was left out, and in the remaining four-fifths of the training dataset a Correlation-Adjusted T (CAT) score was estimated (i.e. a multivariate generalization of the standard univariate T-test statistic that takes the correlation among variables explicitly into account (20) (21)) and the Local False Discovery Rate (LFDR) (i.e. the probability of a variable to be non-informative with regard to phenotype prediction given its CAT score) for each potential predictor. We selected predictors that had a LFDR smaller than 0.8 in > 50% of the rounds (22). This process reduces dimensionality and select variables with higher probability of being informative, reducing the risk of overfitting. These predictors were used to develop predictive models in the training sample using a gradient boosting machine (GBM) algorithm with a five-fold cross-validation repeated 100 and 20 times when predictors were pathway and gene scores, respectively. Cross-validation in this phase was used to provide better estimates of predictor contribution and empirically estimate model parameters (number of trees and interaction depth; shrinkage was set to 0.1 and minimum number of observations in each terminal node was set to 10). GBM produces a prediction model in the form of an ensemble of weak prediction models based on decision trees and it was demonstrated to be a suitable algorithm to learn from weak predictors, when there is not a large amount of available data for training and predictors may interact among each other (23) (24). Models using gene-based scores as predictors included both rare and common variants, because the inclusion of rare variants only would have created scores very skewed towards zero which could not be realistically adjusted for confounders, while models using gene-set scores were tested for rare variants only and rare combined with common variants.

The performance of the developed models in predicting TRD or non-response in the testing sample was estimated using the area under the curve (AUC) of ROC (receiver operating characteristic) curves. Predictive models were developed in the whole training sample and in the subsamples treated with serotonergic antidepressants and noradrenergic antidepressants (only the current treatment was considered, see Supplementary Table 1 for number of subjects), since different genetic profiles were previously reported for these two drug classes (22). The addition of a clinical risk score to the genetic predictors was evaluated. The clinical risk score was calculated as a weighted sum of the variables independently associated with TRD or non-response in the training sample in a regression model after Bonferroni correction (Supplementary Table 3). Each variable included in the clinical score was weighted for its effect size (z score) and divided by the number of variables available in each subject 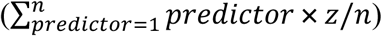 in order to avoid the exclusion of subjects with some missing values. The risk of TRD or non-response may increase particularly at the extremes of the genetic score distribution. Thus, we also tested the significant models including only subjects with a genetic score ≤30 or ≥ 70 percentiles; we used this threshold to balance the risk of instability of findings due to the limited sample size, particularly in the subsamples treated with specific drug classes. The total genetic score was calculated in each subject as a sum of the gene/pathway scores included in the model of interest, each of them weighted for its importance in the predictive model. This approach is a simplification, since it does not reflect the non-linearity of the developed models and possible interactions.

We did not perform multiple-testing correction for these analyses because: 1) these tests were correlated among each other and not independent (for example, patients in the tails of the genetic score are a subset); 2) we looked at the consistency of results of correlated analyses (i.e. we analysed patients in the tails or added the clinical score for further testing models which showed non-random prediction in the basic test).

The following R cran packages were used for the described analyses: caret, nnet, sda, crossval, pROC.

### 2.5. Replication

Replication of the significant predictive models was tested in STAR*D and GENDEP (25) (26), using the same approach described for creating gene- and pathway-based risk scores (including rare and common variants according to the definition reported in paragraph 2.4.2, more details are in Supplementary Methods). In replication samples we used a genetic score ≤20 or ≥ 80 percentiles to identify subjects with extreme genetic scores since the larger sample size. In both these samples genome-wide genotyping was available, including standard genome-wide arrays and an exome array (Illumina Infinium Exome-24 v1.0 BeadChip) (27), but not exome sequence data. Further information on genotyping methods and quality control was previously reported (28) and it is described also in the Supplementary Methods. Imputation was carried out using the Michigan imputation server and the Haplotype Reference Consortium (HRC, version r1.1 2016) as reference panel (29). Different imputation quality thresholds were used to prune rare and common variants according to the previous literature (R^2^>0.30 and R^2^>0.60 for common and rare variants, respectively (30)(31)). The comparability between the available rare variants in GENDEP/STAR*D and GSRD was tested in terms of number and functional annotation. Phenotypes were defined in a way comparable to the GSRD sample (Supplementary Methods).

### 2.6. Power estimation

GSRD sample size after quality control (n=1209) provides adequate power (≥0.80) in 865 out of 1000 simulations when testing a combination of 45 simulated rare variants (MAF < 0.02) and 100 simulated common variants (which reflects the median number of variants in the analysed genes), having effect sizes (β) randomly distributed between -0.25 and 0.25, at alpha=2.69e-06 (Bonferroni corrected p-value for number of genes). R cran libraries KATSP, minqa and CompQuadForm were used for power estimation (32).

## 3. Results

The number of subjects available after quality control was 1209 (details on number of excluded subjects are in Supplementary Figure 2). A comprehensive overview of clinical-demographic characteristics of the samples is reported in Supplementary Table 1, while a condensed overview is shown in Table 1. The number of included variants split by variant type and MAF is reported in Supplementary Table 2 (exome sequence data). Five subjects showed low concordance between genotypes available in both exome and genome-wide data and they were excluded from the analyses including both rare and common variants, since exome sequencing repeated on one of these subjects demonstrated genotype concordance >99% with the initial sequencing results. The comparison between sequenced rare variants and rare variants imputed from genome-wide data showed a mean concordance of 75% (SD=5%) considering only non-major homozygote genotypes. The mean concordance considering the same comparison but for genotyped rare variants (array data) was 93% (SD=2%) (Supplementary Figure 3), suggesting that the use of rare variants obtained from an array may be feasible even though not optimal. From the genome-wide data, 476,319 intragenic common variants in low LD and 1180 subjects were included after quality control.

**Table 1:** main clinical-demographic characteristics of the training sample (n=847) and testing sample (n=362). The baseline MADRS score is referred to the beginning of the current depressive episode. MADRS=Montgomery and Åsberg Depression Rating Scale. TRD=treatment-resistant depression. Mean ± standard deviation is reported for continuous variables and distribution for dichotomous ones. For a more comprehensive overview of patients’ characteristics and results of comparisons between the characteristics of the two subsamples see Supplementary Table 1.

| Variable | Training sample (n=847) | Testing sample (n=362) |
| --- | --- | --- |
| Age | 51.44±13.94 | 51.87±14.16 |
| Gender (F/M) | 566/281 | 235/127 |
| Phenotype of interest | TRD n=353<br>Non-Responders n=291<br>Responders=203 | TRD=151<br>Non-responders n=125<br>Responders=86 |
| Baseline MADRS score | 34.56±7.36 | 33.85±7.69 |
| Current MADRS score | 24.73±11.13 | 24.78±11.60 |
| Treatment | Serotonergic n=421<br>Noradrenergic n=271<br>Serotonergic-noradrenergic n=128<br>Other n=27 | Serotonergic n=192<br>Noradrenergic n=93<br>Serotonergic-noradrenergic n=59<br>Other n=18 |

The variables included in the clinical risk score were suicidal risk, number of previous depressive episodes, chronic depression and two MADRS factors (pessimism and interest-activity) (Supplementary Table 3).

### 3.1. Distribution of damaging variants

Patients with TRD and non-responders did not show an increased risk to carry SIFT/PolyPhen damaging variants compared to responders or variants with SO functional score ≥ 0.90 or ≥ 0.70 (Supplementary Table 4 and Figure 1). When considering individual genes (Supplementary Tables 5-6), we did not identify any difference among phenotypic groups after Bonferroni correction. The top gene was *WDR90* (WD Repeat Domain 90) which showed variants with SO functional score ≥ 0.9 in 21 patients with TRD but only in 4 non-responders and 2 responders (p=3.44e-05).

**Figure 1:**
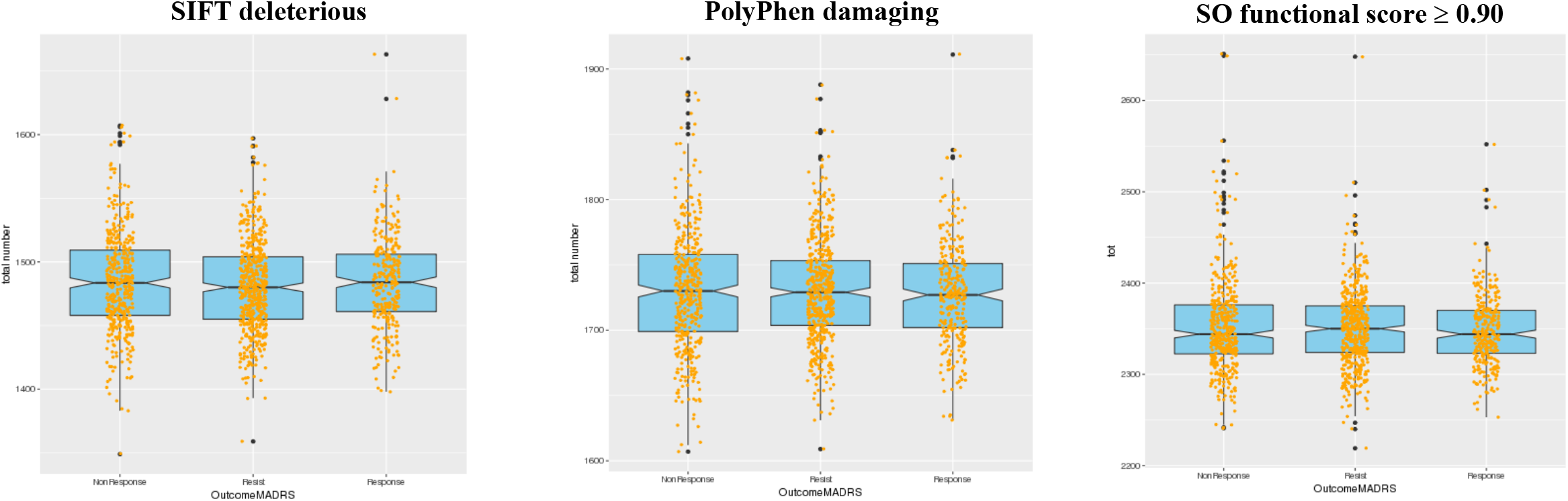
representation of exome-wide distribution of variants with SO functional score ≥ 0.90, SIFT deleterious/deleterious low confidence variants (scores >0.95), PolyPhen damaging/probably damaging variants (scores>0.15). The examined phenotypic groups (x axis) were treatment-resistant depression (TRD), non-response and response. The number of variants in each phenotypic group is reported on the y axis.

### 3.2. Exome-wide, gene and pathway scores

The distribution of the of exome-wide scores for the three tested functional weights were substantially overlapping. Six patients were excluded from the subsequent analyses as they scored outside five standard deviations from the sample mean (Supplementary Figure 4). Patients with TRD and non-responders did not show higher exome-wide scores compared to responders (p>0.05 for all three tested functional weights). The correlations between gene scores calculated using the three tested functional weights were high (mean correlation coefficient between 0.89 and 0.95 with SD from 0.04 to 0.06 in pair-wise comparisons, Supplementary Figure 5). In consideration of these high correlation coefficients, the demonstration that Eigen scores have better discriminatory ability using disease-associated and putatively benign variants from published studies compared to CADD scores (19), and the lower functional precision of SO functional scores, only Eigen-based functional weighting was used in subsequent analysis.

Gene- and pathway-based scores were not associated with phenotypic groups after Bonferroni correction (Supplementary Tables 7-8). The top genes were *NBN* and *ZNF418* (p=4.34e-05 and 5.18e-05, respectively, whole sample, Supplementary Table 4) and the top pathways were PID (protein interaction database) CD40 pathway in the subsample treated with serotonergic drugs and GO (gene ontology) response to cocaine in the subsample treated noradrenergic drugs (p=5.28e-05 and 5.61e-05, respectively, Supplementary Table 8).

### 3.3. Predictive modeling

Pathway-based models for TRD vs. response in the whole sample including only rare genetic variants showed non-random prediction in the testing sample (AUC 0.61 [95% CI 0.54-0.69], Table 2 and Figure 2) and in patients treated with serotonergic antidepressants (n=272 and n=118 in the training and testing samples respectively, AUC 0.62 [95% CI 0.52-0.73], Table 2 and Figure 2). The list of pathways used as predictors is in Supplementary Table 9. No significant prediction of TRD vs. response was observed in the subsample treated with noradrenergic antidepressants or when comparing non-responders vs. responders or TRD plus non-responders vs. responders (Supplementary Table 10). Prediction was improved by adding the clinical risk score to genetic predictors in both the whole sample and patients treated with serotonergic antidepressants (AUC 0.73 [0.66-0.79] and AUC 0.65 [0.55-0.76], respectively, Table 2 and Figure 2), and this effect was more evident in subjects having extreme genetic scores for the included pathways (AUC 0.75 [0.67-0.83] and AUC 0.68 [0.55-0.82], respectively; Table 2 and Figure 2).

**Table 2:**
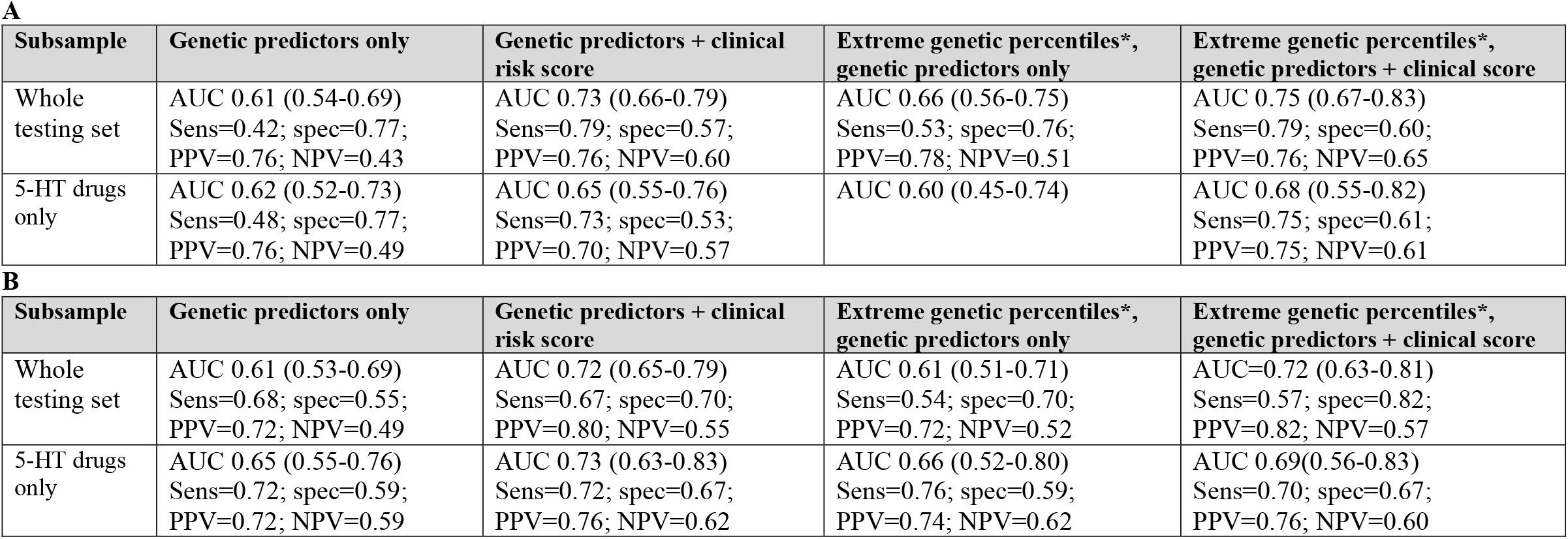
characteristics of the models showing significant prediction in the testing sample for the phenotype TRD vs. response. The results of the other tested models are shown in Supplementary Table 9. **A**. Pathway-based scores including rare variants; **B**. Gene-based scores including common and rare vairants. Sens=sensitivity; spec=specificity; PPV=positive predictive value; NPV=negative predictive value. 5-HT=serotonergic drugs. AUC 95% confidence intervals are reported within parenthesis. * including only subjects with a genetic score ≤30 percentile or ≥ 70 percentile see paragraph

| **A** |  |  |  |  |
| --- | --- | --- | --- | --- |
| Subsample | Genetic predictors only | Genetic predictors + clinical risk score | Extreme genetic percentiles*, genetic predictors only | Extreme genetic percentiles*, genetic predictors + clinical score |
| Whole testing set | AUC 0.61 (0.54-0.69)<br>Sens=0.42; spec=0.77;<br>PPV=0.76; NPV=0.43 | AUC 0.73 (0.66-0.79)<br>Sens=0.79; spec=0.57;<br>PPV=0.76; NPV=0.60 | AUC 0.66 (0.56-0.75)<br>Sens=0.53; spec=0.76;<br>PPV=0.78; NPV=0.51 | AUC 0.75 (0.67-0.83)<br>Sens=0.79; spec=0.60;<br>PPV=0.76; NPV=0.65 |
| 5-HT drugs only | AUC 0.62 (0.52-0.73)<br>Sens=0.48; spec=0.77;<br>PPV=0.76; NPV=0.49 | AUC 0.65 (0.55-0.76)<br>Sens=0.73; spec=0.53;<br>PPV=0.70; NPV=0.57 | AUC 0.60 (0.45-0.74) | AUC 0.68 (0.55-0.82)<br>Sens=0.75; spec=0.61;<br>PPV=0.75; NPV=0.61 |
| **B** |  |  |  |  |
| Subsample | Genetic predictors only | Genetic predictors + clinical risk score | Extreme genetic percentiles*, genetic predictors only | Extreme genetic percentiles*, genetic predictors + clinical score |
| Whole testing set | AUC 0.61 (0.53-0.69)<br>Sens=0.68; spec=0.55;<br>PPV=0.72; NPV=0.49 | AUC 0.72 (0.65-0.79)<br>Sens=0.67; spec=0.70;<br>PPV=0.80; NPV=0.55 | AUC 0.61 (0.51-0.71)<br>Sens=0.54; spec=0.70;<br>PPV=0.72; NPV=0.52 | AUC=0.72 (0.63-0.81)<br>Sens=0.57; spec=0.82;<br>PPV=0.82; NPV=0.57 |
| 5-HT drugs only | AUC 0.65 (0.55-0.76)<br>Sens=0.72; spec=0.59;<br>PPV=0.72; NPV=0.59 | AUC 0.73 (0.63-0.83)<br>Sens=0.72; spec=0.67;<br>PPV=0.76; NPV=0.62 | AUC 0.66 (0.52-0.80)<br>Sens=0.76; spec=0.59;<br>PPV=0.74; NPV=0.62 | AUC 0.69(0.56-0.83)<br>Sens=0.70; spec=0.67;<br>PPV=0.76; NPV=0.60 |

**Figure 2:**
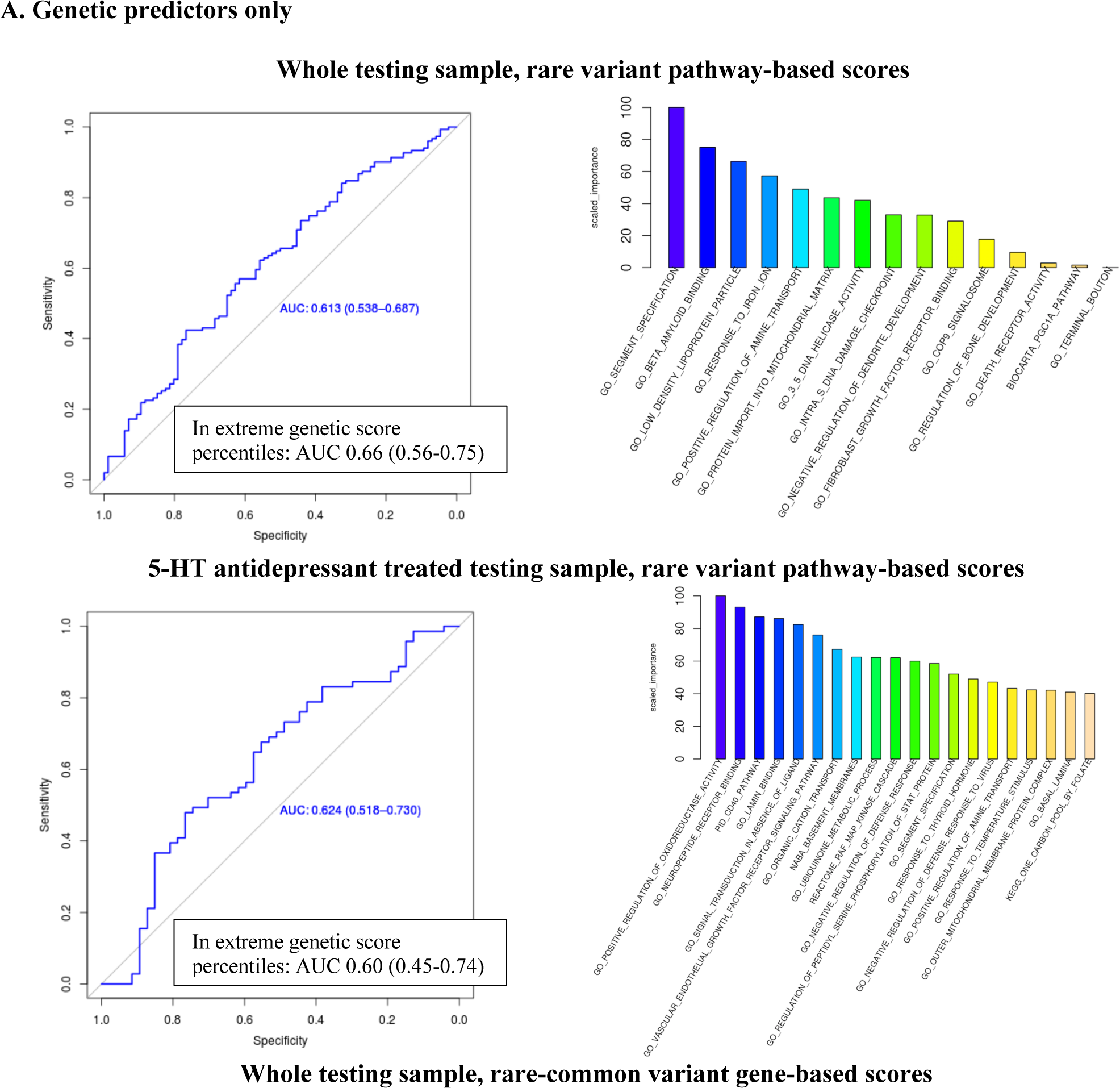
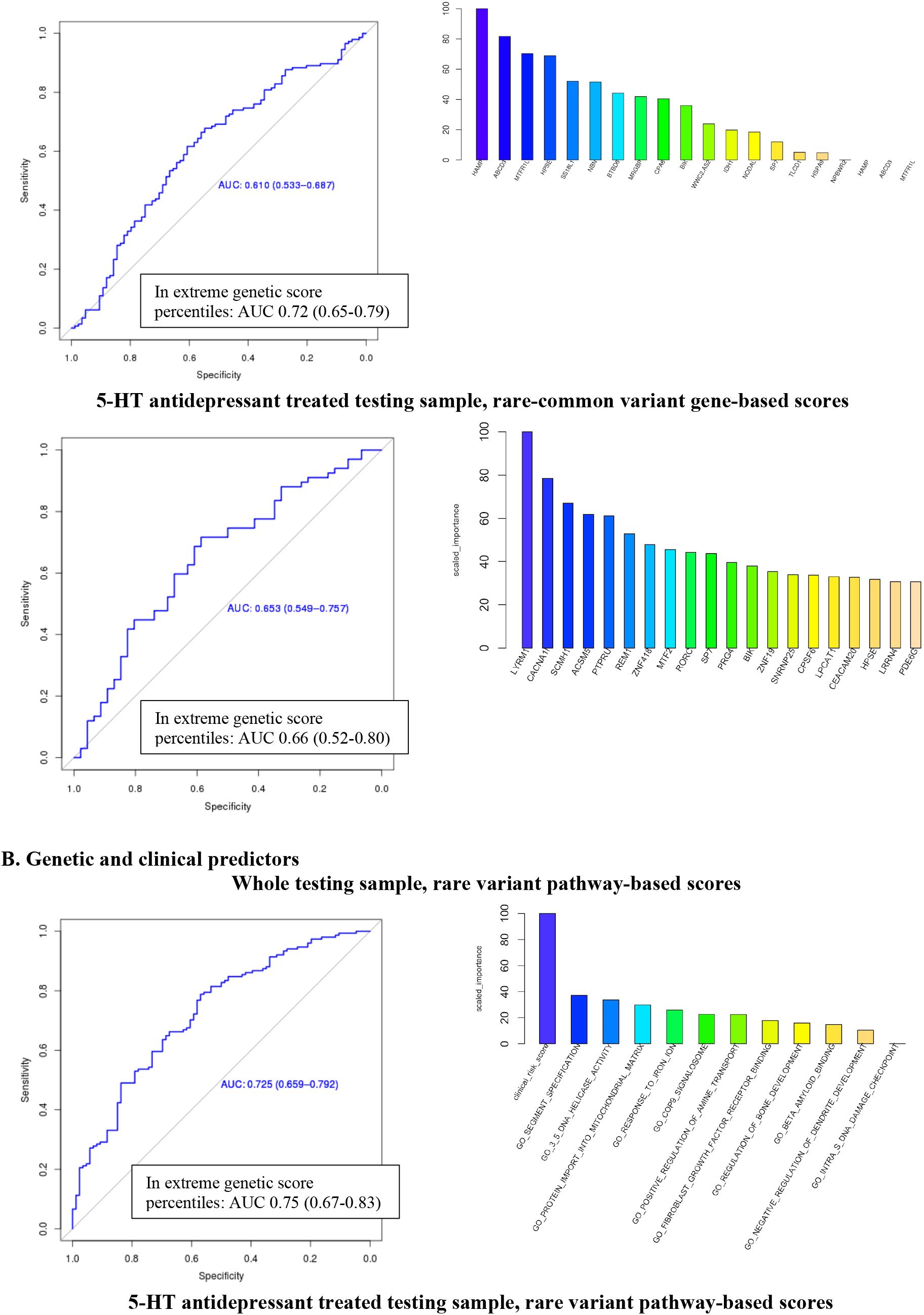
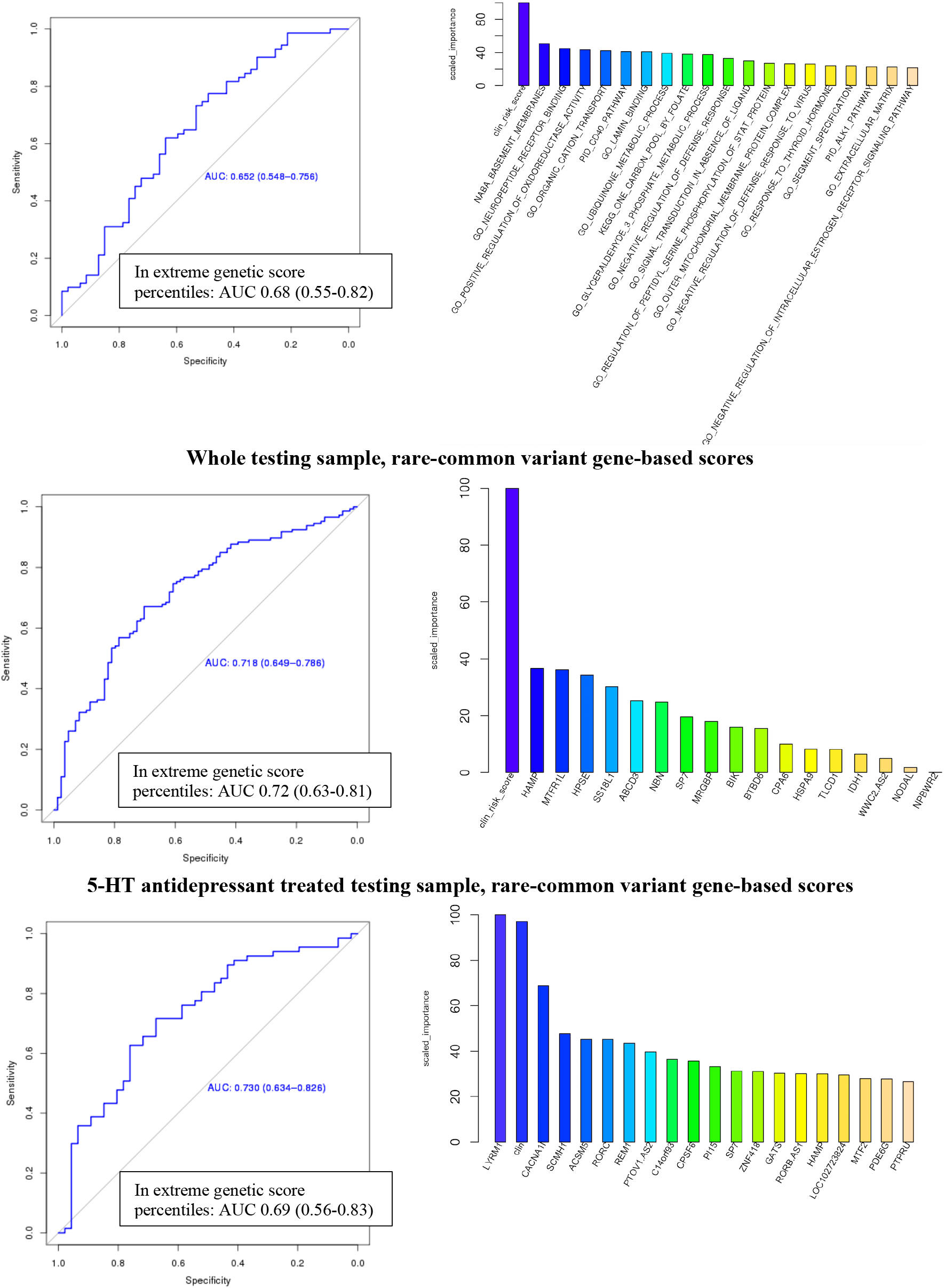
ROC curves of the non-random predictive models in GSRD testing sample and relative importance of the genetic predictors included in the models. When more than 20 predictors were included, only the first 20 are shown. 5-HT=serotonergic. The AUC values reached including only subjects with genetic scores ≤30 or ≥ 70 percentiles. **A**. Genetic predictors only. **B**. Genetic and clinical predictors.

Pathway-based models including rare and common genetic variants did not show predictive effect in the testing sample in almost all scenarios (Supplementary Table 10).

Gene-based models including rare and common variants predicted TRD vs. response in the whole testing sample and in subjects treated with serotonergic antidepressants (AUC 0.61, [0.53-0.69] and AUC 0.65, [0.55-0.76], respectively; Table 2). The lists of genes used as predictors is shown in Supplementary Table 9. The addition of the clinical risk score improved the prediction while the subgroups having scores in the extreme percentiles did not show substantially different results (Table 2). Predictive models of non-response vs. response showed marginal significance in the whole sample (AUC 0.59 [0.51-0.67]) but better values in the sample treated with serotonergic antidepressants (AUC 0.64, [0.53-0.74]; Supplementary Table 10). However, given that models including non-responders were significant in a smaller number of scenarios compared to those focused on TRD, we did not further investigate them.

### 3.5. Replication in STAR*D and GENDEP

Despite the availability of genotypes from an exome array, a low covering of coding regions was obtained compared to exome sequence data, limiting the comparability of these data with those available in GSRD (Supplementary Figure 6). The number of subjects included and their main clinical-demographic characteristics are reported in Supplementary Table 11.

The rare variant pathway-based model developed in the whole GSRD training sample did not show significant prediction in STAR*D or GENDEP, but the addition of the clinical score improved prediction which reached significance in GENDEP (AUC 0.60 [0.54-0.65]). The model developed in subjects treated with serotonergic drugs did not show significant results either, but the results improved when considering subjects at the extremes of the genetic score distribution (≤20 or ≥ 80 percentiles), particularly in STAR*D when adding clinical predictors (AUC 0.61 [0.51-0.71]). Given the larger sample size of STAR*D, we also looked at more extreme percentiles and prediction clearly improved in subjects with scores ≤10 or ≥ 90 percentiles (n=134; AUC 0.73, 95% CI 0.61-0.86). The rare-common gene-based model developed in the whole GSRD training sample showed a weak prediction in STAR*D when the clinical risk score was also included (AUC 0.55 [0.51-0.59]), which however was not obaserved in GENDEP (AUC 0.54 [0.48-0.61]). The model developed in subjects treated with serotonergic drugs was close to significance in both samples and it showed non-random prediction in GENDEP when considering also the clinical factors (AUC 0.62 [0.53-0.72]). This last model was developed in GSRD training set after the exclusion of LCE1B that was not covered in GENDEP, with no major change in predictive performance in the GSRD testing sample (not shown). An overview of the replication results is provided in Supplementary Table 12.

## 4. Discussion

This study found no overall difference in the distribution of functional and deleterious/damaging variants between TRD patients, non-responders and responders within the whole exome or within individual genes. The closest gene to the significance threshold was *WDR90* (WD Repeat Domain 90), which product function is still poorly known but it is thought to participate in microtubule organization and it has probably a role within the presynaptic axon terminal (33). The tested risk scores were not associated with TRD at gene or gene set level, with *NBN* (nibrin) and *ZNF418* (Zinc Finger Protein 418) genes, PID CD40 and GO response to cocaine pathways as top results. *NBN* is thought to be involved in DNA double-strand break repair, DNA damage-induced checkpoint activation and telomere integrity (34). It may be involved in neurodegenerative disorders (35). Variants in the *ZNF418* region had a non-significant trend of association with MDD in a previous PGC (Psychiatric Genetic Consortium) mega-analysis (36) and in an exome sequence study (37). The PID CD40 gene set includes 31 genes, it is involved in the modulation of inflammation and CD40 ligand has been previously associated with MDD (38).

The lack of strong signals coming from individual genes or pathways was expected as it is in line with a previous genome-wide association study of copy number variants (CNVs) that reported no significant enrichment of CNVs in TRD (39). Thus, it is reasonable to hypothesize that if genetics contributes to TRD, as suggested by formal genetic studies, multiple genes/pathways must be involved with complex interactions. This mirrors the highly polygenic liability to MDD that is emerging (40). On the basis of this hypothesis, we applied predictive modeling to assess TRD risk using gene- and pathway genetic as well as clinical scores as predictors. Predictive modeling combining genetic and clinical predictors has been used by only two previous studies to predict antidepressant response to the best of our knowledge (22) (41), both these studies used SNPs from genome-wide genotyping as genetic predictors. In contrast to the present study, they did not perform any independent replication and the second study did not distinguish between training and testing sets (41). The present study applied an innovative approach which combined gene and pathway polymorphisms in genetic scores weighted by their functional relevance, using exome sequence and genome-wide data. The predictive models comparing TRD vs. response showed significant prediction in a higher number of scenarios compared to models testing non-response vs. response or TRD + non-response vs. response, confirming the biological relevance of TRD as a distinct phenotype. In this regard, it should be noted that non-responders are a more heterogeneous group than TRD patients, because part of them is expected to develop TRD. In the GSRD testing sample, both gene- and pathway-based models showed significant prediction of TRD vs. response (Table 2). The genes/pathways included in these models (Supplementary Table 9) are mostly involved in cell survival, cell growth and replication, cell migration, neurodegenerative processes, neuroplasticity, immune system, hormonal regulation (sex and thyroid hormones) and second messenger cascades. Predictive performance was often improved by adding clinical risk factors and in the extreme percentiles of the score distribution, suggesting that clinical applications may be more feasible in subjects with extremely low or high genetic risk factors (40). Predictive performance was 3-6% better (whole sample and serotonergic drugs treated sample, respectively) than when including only clinical risk factors. The addition of clinical variables had a smaller effect in the subset treated with serotonergic antidepressants, because these patients had less frequent clinical risk factors (treatment prescription was naturalistic and serotonergic antidepressants, mostly represented by SSRIs, are typically prescribed to patients having lower clinical risk of non-response). This means that the different gene/pathways selected in the whole sample compared to those selected in patients treated with serotonergic antidepressants may reflect their different clinical characteristics more than differences due to distinctive biological mechanisms implicated in response to different drug classes. None of the analyses performed in the group treated with noradrenergic drugs was significant, a probable consequence of the small size of this group. In this regard, we also underline that polypharmacy was frequent in this sample, including combination and augmentation strategies (10), and most of the participants already had two or more pharmacotherapies during the present depressive episode, thus our classification according to the antidepressant class represented a simplified approach.

The significant models developed in the GSRD testing sample were only partially replicated in STAR*D and GENDEP, and this may be explained by the poor comparability of the available genetic data (only arrays, with low coverage of coding regions). The models showing the best performance were the ones developed in subjects treated with serotonergic antidepressants. These were the models showing the greatest effects of genetic predictors and an improvement of prediction when considering extreme percentiles of the genetic predictors. The greatest effect was found for pathway-based genetic scores in STAR*D subjects treated with serotonergic antidepressants at the extremes of the score distribution (AUC 0.73, [0.61-0.86]). The results in the replication samples, despite the low comparability with GSRD, suggest that genetic factors associated with resistance to serotonergic antidepressants may be distinctive, in line with a previous study (22). However, we underline that the total score distribution used to identify subjects with extreme genetic risk factors does not reflect the non-linear modeling used and it may therefore be an oversimplification. In addition to the low comparability of genetic data in the replication samples, there were also relevant clinical differences between STAR*D, GENDEP and GSRD. For example, patients in STAR*D had very long depression of relatively mild severity on average and a relatively high number of previous depressive episodes, while in GENDEP there were no patients with chronic MDD according to the standard definition (>=2 years) and they had on average a lower number of previous episodes (Supplementary Table 11). Unlike the other samples, MADRS was not available in STAR*D and equivalent scores were calculated using the QIDS-C16 scale. In GENDEP the definition of TRD was based on retrospective data which were not collected for this purpose, potentially resulting in phenotyping errors. The combination of the clinical risk score with genetic predictors was not always straightforward, because of the possible dynamic interactions between them. This may explain why we obtained a worse prediction including the clinical risk score compared to genetic predictors only for some of the tested models. The approach used to create the clinical risk score aimed to avoid the exclusion of subjects with partially missing data, but it provides a cumulative estimation and the contribution of the individual clinical risk variables could not be considered in the predictive models. Finally, our predictive models do not show suitable performance for clinical application, particularly because of relatively low specificity (we were able to identify TRD patients in a quite satisfying way, but not responders).

Bearing in mind the discussed limitations of this study, our results contributed to clarify the genetic factors involved in non-response to treatment in MDD and TRD. This is also the first study to assess the contribution of rare genetic variants to these traits through whole exome sequencing, if we exclude a very small pilot study performed on 10 subjects (9). No individual gene or pathway probably plays a major role in TRD, thus models including multiple genes/pathways and able to account for their interactions are probably the best strategy. Theoretically, pathway-based models are more suitable to take into account the complex genetic component of antidepressant response compared to gene-based models and they are expected to be more replicable, as confirmed by our top replication result. Our study represents a new approach to the prediction of treatment response and resistance in MDD and future improvements may lead to clinical applications, at least in patients with extreme genetic scores. In patients having genetic risk for TRD, treatment strategies with demonstrated higher efficacy (e.g. pharmacotherapy combined with psychotherapy (42)) but limited availability for cost constraints could be implemented as first line treatment, when these patients first seek treatment and there are still no clinical signs of severe MDD, reducing the proportion of patients at risk who progresses towards resistance.

## Data Availability

Individual-level data or summary statistics is not publicly available for GSRD and GENDEP, however raw genotypes and phenotypes of these samples are available to Psychiatric Genomic Consortium (PGC) analysts (https://www.med.unc.edu/pgc/) within approved projects. Individual-level data is publicly available for STAR*D through the NIMH genetics website (https://www.nimhgenetics.org), after submission of a research project and access authorization.

## Acknowledgements

We thank the NIMH for having had the possibility of analyzing their data on the STAR*D sample. We also thank the authors of previous publications in this dataset, and foremost, we thank the patients and their families who accepted to be enrolled in the study. Data and biomaterials were obtained from the limited access datasets distributed from the NIH-supported “Sequenced Treatment Alternatives to Relieve Depression” (STAR*D). The study was supported by NIMH Contract No. N01MH90003 to the University of Texas Southwestern Medical Center. The ClinicalTrials.gov identifier is NCT00021528.

The GENDEP project was supported by a European Commission Framework 6 grant (contract reference: LSHB-CT-2003-503428). The Medical Research Council, United Kingdom, and GlaxoSmithKline (G0701420) provided support for genotyping. This paper represents independent research part-funded by the National Institute for Health Research (NIHR) Biomedical Research Centre at South London and Maudsley NHS Foundation Trust and King’s College London. The views expressed are those of the author(s) and not necessarily those of the NHS, the NIHR or the Department of Health and Social Care. High performance computing facilities were funded with capital equipment grants from the GSTT Charity (TR130505) and Maudsley Charity (980).

We thank Intomics (Copenhagen, Denmark) for genotype calling and contribution to quality control of exome sequence data in the GSRD sample.

